# Challenges and best practices for digital unstructured data enrichment in health research: a systematic narrative review

**DOI:** 10.1101/2022.07.28.22278137

**Authors:** Jana Sedlakova, Paola Daniore, Andrea Horn Wintsch, Markus Wolf, Mina Stanikic, Christina Haag, Chloé Sieber, Gerold Schneider, Kaspar Staub, Dominik Alois Ettlin, Oliver Grübner, Fabio Rinaldi, Viktor von Wyl, University of Zurich Digital Society Initiative (UZH-DSI) Health Community

**Author notes:** Corresponding Author: Prof Viktor von Wyl, Institute for Implementation Science in Health Care, University of Zurich, 8006 Zurich, Switzerland.

## Abstract

Digital data play an increasingly important role in advancing medical research and care. However, most digital data in healthcare are in an unstructured and often not readily accessible format for research. Specifically, unstructured data are available in a non-standardized format and require substantial preprocessing and feature extraction to translate them to meaningful insights. This might hinder their potential to advance health research, prevention, and patient care delivery, as these processes are resource intensive and connected with unresolved challenges. These challenges might prevent enrichment of structured evidence bases with relevant unstructured data, which we refer to as digital unstructured data enrichment. While prevalent challenges associated with unstructured data in health research are widely reported across literature, a comprehensive interdisciplinary summary of such challenges and possible solutions to facilitate their use in combination with existing data sources is missing.

In this study, we report findings from a systematic narrative review on the seven most prevalent challenge areas connected with the digital unstructured data enrichment in the fields of cardiology, neurology and mental health along with possible solutions to address these challenges. Building on these findings, we compiled a checklist following the standard data flow in a research study to contribute to the limited available systematic guidance on digital unstructured data enrichment. This proposed checklist offers support in early planning and feasibility assessments for health research combining unstructured data with existing data sources. Finally, the sparsity and heterogeneity of unstructured data enrichment methods in our review call for a more systematic reporting of such methods to achieve greater reproducibility.

## Introduction

Digitalization has given access to a broad variety of digital unstructured data that contain health-relevant information and can substantially contribute to health research. Digital data in healthcare originate from a wide array of sources, ranging from structured clinical data, such as laboratory test results or patient-reported outcome measures, to unstructured data, such as free text data, collected within or outside of a clinical setting.^1^ This wealth of data holds great potential to advance health research, prevention, and patient care delivery. However, over 80% of digital health data is available as unstructured data,^1^ requiring new forms of data processing and standardizing that prove challenging to health researchers. The challenging nature of digital unstructured data is also reflected in the fact that these data are often not specifically collected for research purposes (e.g., data from social media).

Unstructured data are commonly defined as data that are not readily available in predefined structured formats such as tabular formats.^2,15,21,27^ However, there is no unified, standardized definition of unstructured data in health research. In the literature, unstructured data are often referred interchangeably as “big data”, “digital data”, “unstructured textual data” and described as “high-dimensional”, “large-scale”, “rich”, “multivariate” or “raw”.^1,3,21,25,26,28^

Unstructured data can be utilized on their own or be combined with other data sources to enable *data enrichment* in health research. In this context, we refer to digital unstructured data enrichment to describe the process of augmenting the available evidence base in health research, which mostly consists of structured data with unstructured data.^4^ For example, open-ended patient self-reports or smartphone data can be used to complement longitudinal laboratory, clinical, and survey data.^20,30,42^ Through digital unstructured data enrichment, further insights into individuals’ lifestyles and behaviors can be gained due to the real-time measurements and monitoring data in a natural living environment, contributing to digital phenotyping^5^ and better understanding of health risks or diseases.^30^ Furthermore, it can enable one to access under-researched population groups (e.g., ethnic minorities)^6^ and to gain a deeper understanding of participants’ daily life contexts over longer time periods, as well as outside of clinical settings.^18^ As such, this wealth of integrated data can foster personalized, adaptive, and just in time health status assessments that can be of greater relevance to the study participants.^7^

While the abundance of digital unstructured data presents opportunities in advancing health research, methodological challenges surrounding their extensive preprocessing requirements for meaningful information extraction and integration persist. ^8,15,19,21,30^ These challenges are accentuated as digital unstructured data are increasingly used to develop AI/ML models on unsupervised approaches, rather than on the standard supervised approaches.^9^ As a result, the established scientific process of creating and testing hypotheses is challenged in such a way that hypotheses are more strongly linked with the available data themselves.^10^ These persisting challenges and methodological developments are currently not addressed in the literature, as available methods mainly inform the pre-processing or optimization of computational possibilities with digital unstructured data, rather than informing health research study planning and conduct. As such, there is a need for guidance based on standards and best practices integrating different disciplines to inform the initial phases of study planning in health research with digital unstructured data.

## Aims

This systematic narrative review aims to explore current standards and requirements to use digital unstructured data and their combination with existing data in health research. Specifically, we aim to answer the following research question:

*How can health researchers enable the proper (systematic, reliable, valid, effective, and ethical) use of digital unstructured data to enrich a knowledge base from available data sources?*

To answer this research question, this review 1) identifies and describes the main challenge areas associated with the use of unstructured data to enable digital unstructured data enrichment in health research; 2) provides a summary of possible solutions for common challenges associated with digital unstructured data enrichment; 3) provides guidance for the initial assessment of whether the inclusion of unstructured data is a feasible and appropriate for the study intended research tasks.

The goal of this review is to inform the planning and implementation surrounding the use of unstructured data in health research to enable knowledge enrichment from a methodological perspective.

## Methodology

### Definitions

We define *unstructured data* in accordance with the literature as raw data that are not in a pre-defined structure (e.g., tables) or data that may be structured, but still require substantial pre-processing or feature extraction effort.^15,19,21,30^ Furthermore, we define *digital unstructured data enrichment* as the use of unstructured data in combination with other data sources to contribute to relevant domain knowledge in health research and clinical practice.

In this review, we consider text data, electronic health records (EHR), sensory data from wearables and other devices, including electroencephalogram (EEG) as common sources of unstructured data. Despite their widespread use in health research, we did not consider imaging data in this review, as these data are often bound to manufacturer-proprietary algorithms, creating specific challenges in the enrichment process that may not generalize to other unstructured data types.

### Search Strategy

We conducted a systematic narrative review guided by the Preferred Reporting Items for Systematic Reviews and Meta-Analyses (PRISMA) 2020 statement.^11^ Our study selection was guided by the inclusion and exclusion criteria displayed in **Textbox 1** and **Textbox 2**, respectively. We performed our search on PubMed and PsycInfo for 1) general overview articles, 2) primary research articles, 3) and articles describing databases, all including relevant information on digital unstructured data enrichment. Our search was restricted to articles from the fields of neurology, cardiology, and mental health. These were chosen due to the high prevalence of unstructured data availability in these fields and their established use for research and healthcare.^12,13^ The complete search syntax including all keywords can be found in **Appendix 1**.

Screening was conducted in two phases. In the first step, we screened the titles and abstracts from the studies based on the inclusion criteria (**Textbox 1**). In the second step, we performed a full-text screening of the articles selected in the first step and excluded articles that matched the criteria outlined in **Textbox 2**. In both steps, one investigator (JS) assessed all articles and a second investigator (PD) performed checks on a randomly selected sample of articles for each screening phase. Any disagreements were discussed and, if required, a decision was achieved through the principal investigator (VvW).

**Textbox 1.**
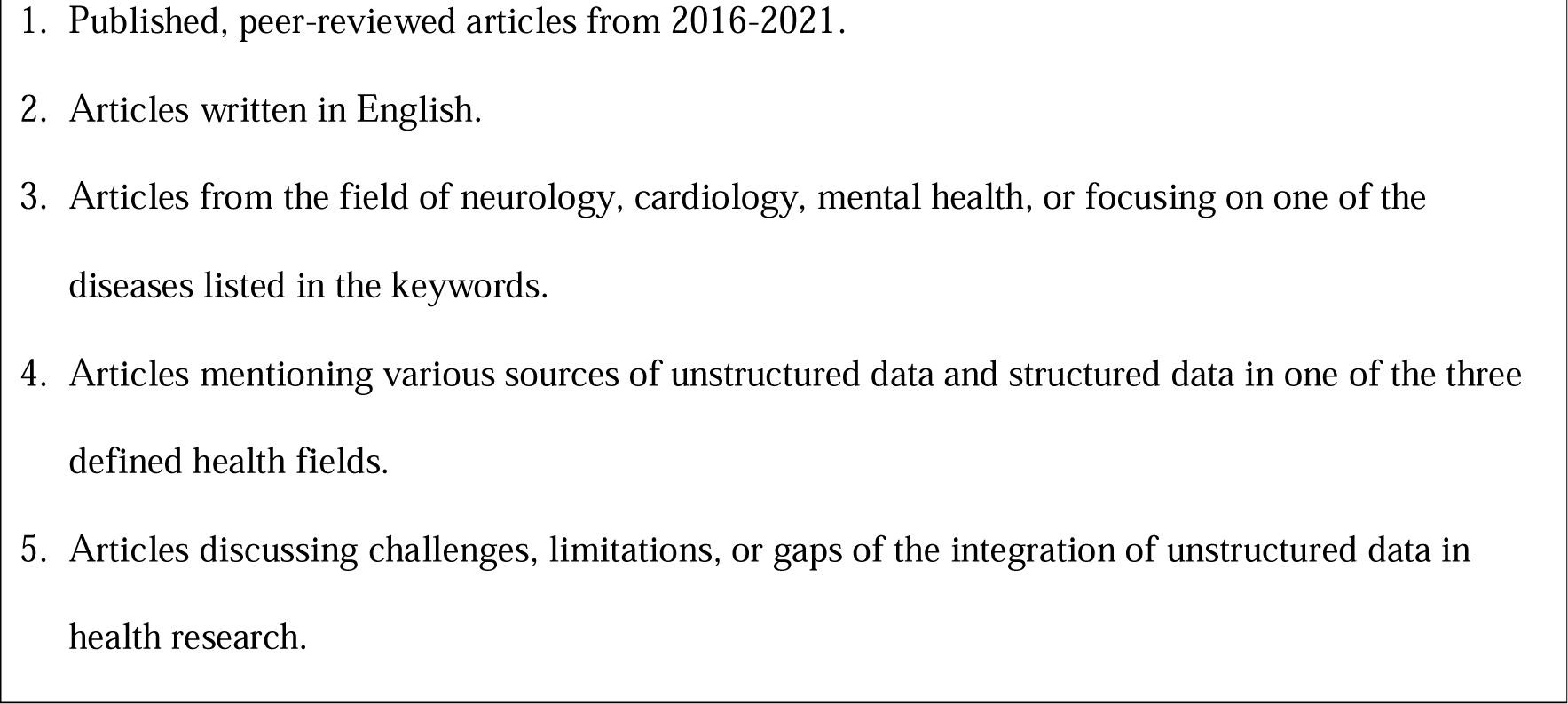
Literature Review Inclusion Criteria

**Textbox 2.**
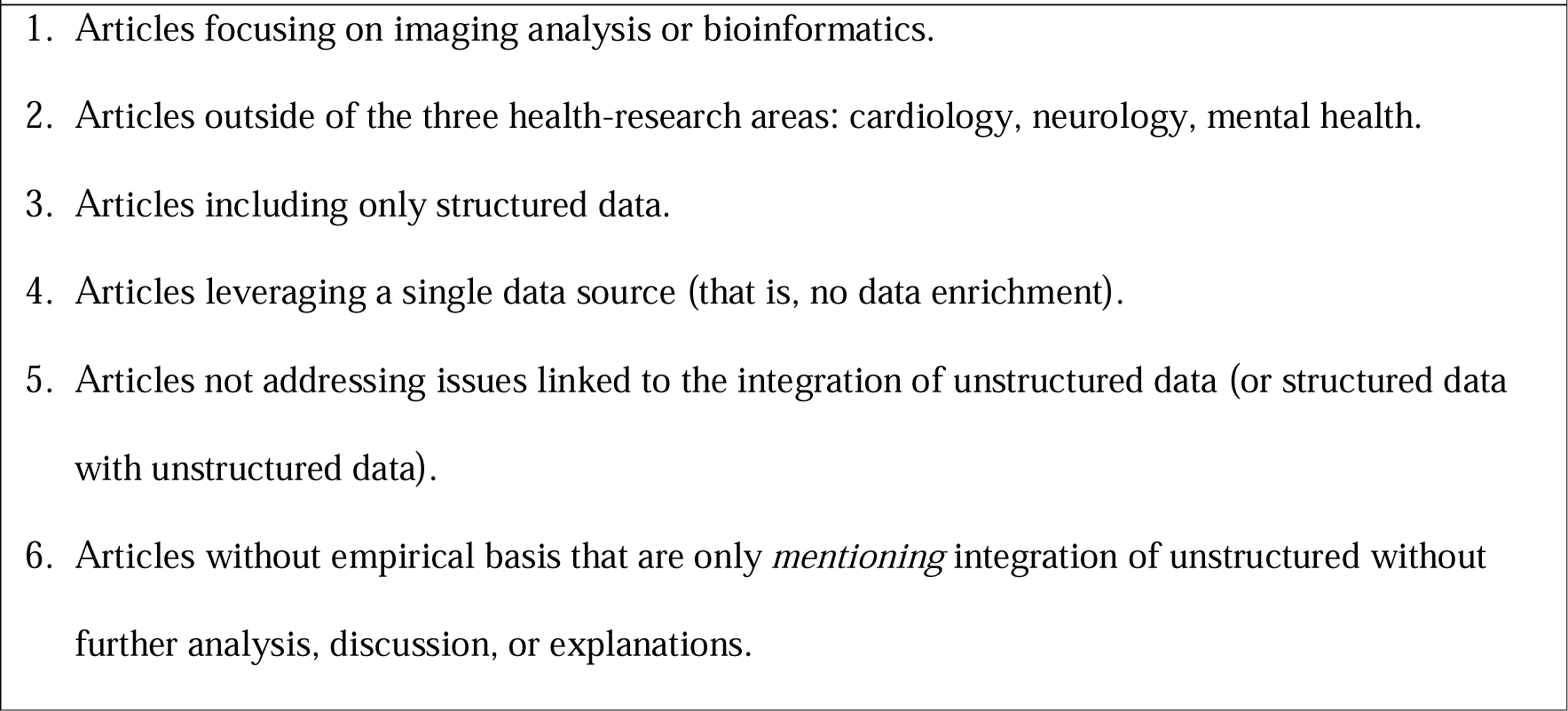
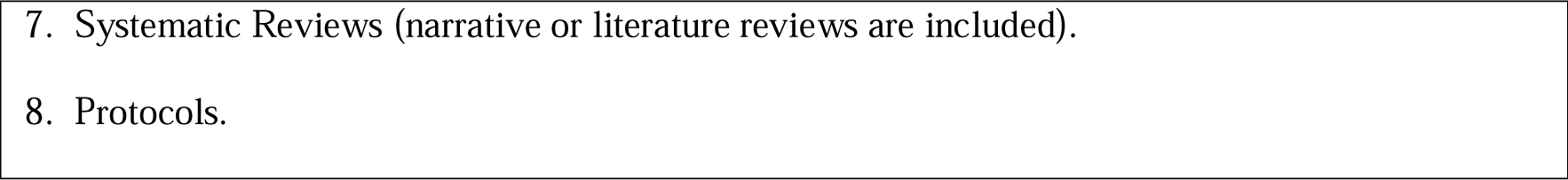
Literature Review Exclusion Criteria

### Data Extraction and Synthesis

Data extraction was standardized yet developed iteratively. The initial data extraction was based on standard study characteristics and guided by our research question. During the full-text screening, seven overarching topics related to digital unstructured data enrichment were identified and used for data extraction. The topics were the following: 1) medical field and subfield of the study, 2) main motivation for unstructured data integration, 3) data enrichment scope (e.g., gathering accurate information about disease severity), 4) type(s) of unstructured data, 5) limitations of unstructured data (e.g., quality/completeness), 6) challenges of data integration, and 7) proposed or discussed approaches for overcoming the mentioned challenges.

A narrative synthesis of the results was conducted to provide an overview on the challenges and proposed solutions related to the digital unstructured data enrichment. This choice was also influenced by the heterogeneity of included studies that ranged from overview papers to original research studies. To address study aims 1 and 2 (i.e., description of common challenges and their possible solutions), the extracted study data on the topics 5 and 6 (i.e., limitations and challenges associated with enabling digital unstructured data enrichment) were grouped into challenge areas. These challenge areas were defined on the basis of major overarching topics 5 and 6 identified after a first full-text screen of the included studies. The challenge areas include not only topics directly connected with data enrichment, but also related to the unstructured data use itself, as this is an essential requirement to enable digital unstructured data enrichment. For each challenge area, relevant possible solutions to tackle the challenges were summarized. For study aim 3 (i.e., providing guidance), we developed a preliminary checklist based on findings from our literature review to guide early study planning and feasibility assessment steps for studies aiming to include unstructured data. To this end, the identified challenge areas from study aim 1 were re-phrased into checklist questions and ordered according to the common study planning stages in health research.^14^ Finally, the checklist was complemented and refined based on domain-specific expertise represented by the interdisciplinary team.

## Results

Our database search yielded 358 articles (**Figure 1**). Overall, 28 articles were included for assessment in this review.

**Figure 1.**
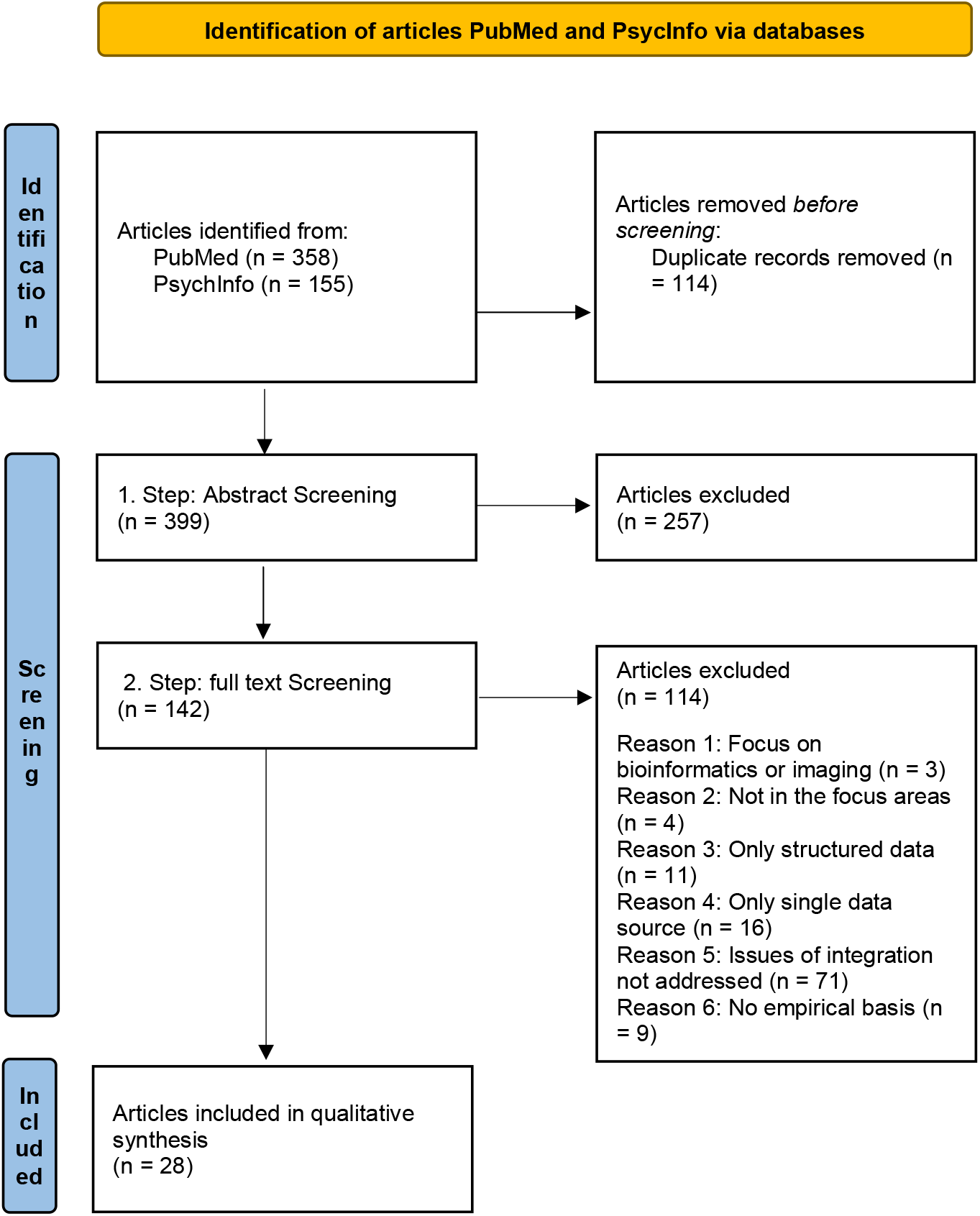
PRISMA Flow Diagram.

### General description of included studies

A description of the 28 included articles^15,16,17,18,19,20,21,22,23,24,25,26,27,28,29,30,31,32,33,34,35,36,37,38,39,40,41,42^ is presented in **Supplementary Table 1** (**Appendix 2)**. The most frequently discussed types of unstructured data sources in the selected articles were electronic health records (n=11) and sensor data (n=7). The most commonly cited motivations for digital unstructured data enrichment were to include of more objective measures in their research, for example, to improve understanding of disease mechanisms and disease prediction, and to strengthen the existing evidence base in precision medicine, real-time monitoring, and real-world data collection.

The most prevalent challenge areas in enabling digital unstructured data enrichment were: 1) the lack of meta-information for unstructured data (n= 6), 2) standardization issues (n= 20) 3) data quality and bias in data (n= 12), 4) infrastructure and human resources (n= 12), 5) finding suitable analysis tools, methods and techniques (n= 14), 6) alignment of unstructured data with a research question and design (n= 11), as well as 7) legal and ethical issues (n= 11). These challenges span across all study stages involving data in a health research study: from data collection to data interpretation. Definitions of the main challenge areas and a brief explanation of their relevance for health research are given in **Supplementary Table 2 (Appendix 2)**.

### Challenge Areas

In the next sections, we summarize the seven identified challenge areas associated with enabling digital unstructured data enrichment in health research and the proposed possible solutions to address them.

#### 1. Lack of meta-information for unstructured data

##### CHALLENGES

Lack of meta-information (e.g., describing data structure and properties or sample population) has been acknowledged as an obstacle for unstructured data findability, integration, interchangeability, and interpretation.^15,19,26,27^ Insufficient meta-information might limit the translation of a study’s findings into clinical practice^15,26^ as important contextual information, such as information on the time in which the data were collected might be missing to assess the usability and correct interpretation of data.^15^

##### POSSIBLE SOLUTIONS

Proposed possible solutions included the standardization of meta-information (e.g., through a standardized format for meta-information through open science standards), which may also resolve issues of data interpretation and their alignment with research questions and designs.^15,19,21,27,35^ Specifically, one suggestion was to provide information for four important aspects in each study: subjects included in the data, context of collection, observations, and time of data collection.^15^ Moreover, a greater availability of standardized meta-information was suggested, as this would facilitate to determine the suitability of specific unstructured data for a given research question.^15,21^

#### 2. Standardization issues

##### CHALLENGES

The most frequently discussed challenge (n= 20) was the lack of a standardized framework for the description of disease phenotypes (e.g., symptoms, clinical presentation), as well as a lack of commonly defined terminologies, ontologies, and data labels.^15,16,17,18,19,21,22,25,26,27,28,29,35,37,40,42^ For example, different terms may be used for a seizure with alteration of consciousness by different physicians^16^ or for the administration of a specific dose of a given drug.^22^ These issues are particularly prevalent in EHRs or clinical annotations^19^ where, for example, terminologies and phenotyping may differ across healthcare settings or change over time.^16,21,23^ There is also an observed lack of standardized data management methods^27,37^ and regulatory standards to guide and assess the use of novel technology and their associated unstructured data in clinical applications such as clinical trials.^15,30^

##### POSSIBLE SOLUTIONS

In most articles, harmonization of data formats, data models, terminologies, ontologies, and analytical tools, as well as working practices were proposed as possible solutions to standardization issues.^15,20,21,22,24,27,28,29^ A consensus of standards across the entire data flow,^15,21,22,27,28^ the effective use of datasets,^27^ data optimization,^29^ data consistency^23,41^ and replicability of the studies^20,21^ were also suggested as a means to foster data sharing. The adoption of unified data standards was considered to be important in both academic and industry settings.^28^

To improve standardization efforts and data sharing, the systematic adoption of FAIR (Findable, Accessible, Interoperable, Reusable) Guiding Principles for scientific data management^43^ was proposed.^28^ Other authors mentioned the need for specialized organizations to promote harmonization of terminologies in health research.^21,28^ An example is the consortium behind the Fast Health Interoperability Resources (FIHR) standard to enable “interoperable communication and information sharing between various healthcare systems”.^28^

#### 3. Data quality and biases in data

##### CHALLENGES

Data quality of unstructured data was frequently cited as an important challenge for evidence creation.^17,19,20,21,23,26,27,29,32^ Unstructured data are often collected for purposes other than research and may lack systematic collection and the rigor of study-based measurements, thus often leading to missing data.^19,20,21,23,26,27,29,39^ In medical records, for example, missing data can occur because health care professionals may omit some information or because of patients’ refusal to share data.^26^ The challenge of data quality is reinforced by data inconsistencies and inaccuracies.^20,21,23,26,29,32^ Other recurrent challenges stem from biases in data collection – mainly in the form of selection and information bias^17,19,23,24,26,27,29^ – and confounding.^21,23^ Selection bias was mentioned, for example, in the context of studies where the sample comprised only of individuals who have the digital literacy skills or interest to share unstructured data from social media or wearable sensors (5, 8). Information bias, such as observer bias, was often mentioned in the context of making errors with data in EHRs use and big data analytics.^17,23,24,26^ Further biases may establish themselves in analyses if processing algorithms were trained on biased data.^19,23,24^ Finally, the quality and continuity of data might be negatively impacted by technical issues that can arise, for example, by software updates of wearable sensors.^27^

##### POSSIBLE SOLUTIONS

Several strategies were proposed for assessing and ensuring data quality.^15,19,21,23,26,27^ For studies that use digital health technologies, one study cited a recommendation from the European Medicines Agency (EMA) urging researchers to define “small, well-defined, meaningful measures followed by a data-driven development path”.^27^ Furthermore, possible data quality issues should be considered for all study phases, including preprocessing, feature extraction or analysis.^19^ First, preprocessing should yield only verified and valid dataset that properly combines the unstructured data with other data sources, for example by ensuring that study samples are representative of the populations that are being studied. Second, following feature extraction, data should be critically assessed for their validity and meaning. Finally, analytical methods should be aligned with the research goals of description, prediction, or prescription of the study in such a way that bias is reduced.

Other studies highlighted the need for a data quality standard checklist such as Data Access Quality and Curation for Observational Research Design (DAQCORD).^19,44^ This checklist should provide a priori guidance for planning of large-scale study data collection and pre-processing,^44^ thereby countering the pervasive practice of post-hoc methods for data cleaning.^19^ Other proposed possible solutions included the use of meta-information to increase data quality, to detect potential biases in the data,^15^ to enable cross-referencing of multiple data sources involving the same individuals, as well as to encourage the comparison of results.^21^ Imputation procedures for addressing missing data, as well as algorithms for checking data quality were also recommended.^19^ Furthermore, the inclusion of study participant feedback can inform data collection and processing and improve the relevance of study findings for the intended target population.^27^

#### 4. Infrastructure and Human Resources

##### CHALLENGES

Several studies pointed out challenges related to infrastructure availability, including databases, or open-source platforms.^15,19,21,22,25,26,27,28^ Infrastructure challenges can be particularly problematic when healthcare data are spread across multiple medical systems that lack connection or interoperability, thus creating isolated data clusters.^22,25,28^ Difficulties in data linkage can also emerge when information system architectures cannot accommodate data standardization and other linkage processing tools.^19^ Furthermore, the lack of skills and formal training opportunities for infrastructure utilization or inadequate knowledge of novel statistical tools and methods for combining unstructured data with other data sources can inhibit their use in health research.^21,26,27^

##### POSSIBLE SOLUTIONS

Improvements such as searchable catalogues, databases and the availability of open platforms can mitigate infrastructure-related challenges.^16,18,19,20,21,24,27,28^ Similarly, the availability of infrastructure for the storage and integration of unstructured data can enable collaborative efforts, facilitate standardization, and foster the alignment of unstructured data with good research question development and research design.^21^ Furthermore, the availability of secure collaborative platforms and repositories for data sharing through open science can enable independent knowledge gain and foster new research studies.^16,27^ Meta-databases or catalogues that facilitate the discovery of open data and linking data across public repositories can also facilitate digital unstructured data enrichment.^21,28^ Several studies further suggested that platforms for integrating datasets from various sources should have a modular, flexible, and scalable structure^16,19,28^ and recommended to define the purpose and goals of such platforms during their development.^20,24^ Open data and open software repositories also provide more opportunities for external validation of novel algorithms or (electronic) clinical outcome measures.^18^ Finally, awareness about novel digital unstructured data enrichment methods, their methodological requirements, and the need for specialized training opportunities should be increased.^20,21^

#### 5. Finding suitable analysis tools, methods, and techniques

##### CHALLENGES

The complexity of analyses and appropriate methodological choices associated with the unstructured data enrichment in health research are challenges that were addressed in multiple studies.^16,19,21,23,24,26,28^ Typical features of unstructured data such as high volume or complexity may be overwhelming for researchers due to a lack of methodological knowledge.^26^ Furthermore, the validity of results may be decreased by algorithms that are either not trained sufficiently or may need recurrent fine-tuning to ensure that they create a model representative of its intended purpose and without biases.^18,19,26^ Working with unstructured data requires specific expertise, typically from data scientists. However, the lack of supply of data scientists or the failure to build effective collaborations with external experts were also cited as impediments to managing the complexity of unstructured data.^21,24,28^ Furthermore, there is a lack of guidelines and standards to guide decisions on which tools, methods, and analytical approaches to use when using unstructured data in health research.^21,26^

We further observed a discrepancy in approaches to reduce the complexity of unstructured data (e.g., using feature extraction) in our studies. While some authors argued that complexity reduction is a feasible and appropriate method to enhance unstructured data integration, others voiced concerns that complexity reduction can also reduce richness of unstructured data – particularly in the context of EHRs.16,42

##### POSSIBLE SOLUTIONS

The complexity of unstructured data calls for increased collaboration among different experts. The increasing need for interdisciplinary efforts among health researchers, data scientists, biostatisticians, and health-care professionals was highlighted by most sources.^15,16,17,18,20,21,27,29^ Some authors emphasized the need for a novel profession that combines expertise in health research and informatics.^21,29^ Many also called for greater attention to trainings of health researchers regarding novel methods for using and combining unstructured data with other data sources.^18,20,21^ The need for specific sets of skills, resources, and guidelines for the successful implementation of big data tools into clinical workflows was further mentioned as a requirement to manage unstructured data complexity.^23^ Furthermore, some authors called for more efforts to develop and establish validated algorithms to process and integrate data.^26^ One suggestion was to “provide AI with more ‘functional’ information, such as domain-specific medical reasoning processes and policies based on heuristic-driven search methods derived from human diagnostician methods”.^28^ In the field of mental health, it was advised to complement data-driven research with qualitative research to strengthen the relevance and meaning of results.^24^

#### 6. Alignment with a research design and/or research question

##### CHALLENGES

The difficulty of finding suitable datasets and their subsequent, critical evaluation for clinical relevance was discussed from several perspectives.^17,18,21,23,24,26,29^ One study strongly warned against adjusting the research agenda to the data that are available.^24^ Furthermore, the fact that unstructured data or technologies generating these data were not designed for scientific purposes^17,18,20,24^ might lead to misinterpretation of the data.^24^ The lack of contextual (meta) information, for example about the data generation process, and observational nature of many sources of unstructured data may limit the value of the data for their use in robust, replicable confirmatory analyses (e.g., regarding disease etiology or intervention).^17,24^ The need for further and robust validation of results or outcomes from unstructured data analyses was a further topic of concern.^20,23,24^ For example, predictive models need further validation before being integrated into clinical settings^23^ and informing clinical decision-making.^26^ Similarly, while linked EHRs are suitable for generating research questions, unstructured data should not be used for influencing clinical practice without prior validation.^21^

##### POSSIBLE SOLUTIONS

It should be ensured that unstructured data are relevant for a research question and desired therapeutic effect.^18^ When working with data from digital health technologies, the EMA recommendation framework - that was developed with the collaboration with industry representatives with the aim to provide insights and guidance on validation and qualification processes of digital technologies^45^ - could be consulted for guidance with research question design.^27^ Another recommendation was to align large-scale research projects using unstructured data with clinical priorities and outcome-focused research.^20^ Similarly, the choice of analytical tools depends on the goals of health research: description, prediction, or prescription.^19^ Thus, setting clear research goals might help with the choice of appropriate analytical tools and methods. Finally, unstructured data should be used rather with complementary and enrichment purposes than as a replacement of other traditional methods or datasets.^18,23,24,30^

#### 7. Ethics & Legal Issues

##### CHALLENGES

The most frequently mentioned ethical challenges concerned privacy protection, informed consent and preservation of individual agency over data use.^19,20,21,23,25,26,27,32^ Further challenges connected with digital unstructured data enrichment include inappropriate patient profiling^23^ and decreased participants diversity due to low digital literacy skills reducing some participants’ contributions to certain types of unstructured data (e.g., from social media use).^27^ Furthermore, current deidentification and anonymization practices may still allow patient-linkage. This is, for example, enabled when a combination of data on unusual physical conditions of a patient from a local hospital or a combination of gender, age and admission date might be unique enough to identify a subject and connect it with consumer-level data.^19,42^

##### POSSIBLE SOLUTIONS

Strategies for preserving data privacy and security were discussed in multiple studies.^18,19,20,21,25,26,27,35^ Some authors proposed to develop a new social contract and a broad consent model to balance the benefits of data usage and privacy concerns.^20,21,26^ Unified rules for data governance across fields and sectors might contribute to systematic privacy protection and confidentiality,^19^ such as through unified procedures for data anonymization. Additionally, the importance of engagement with regulatory agencies in early stages of research was emphasized to ensure alignment of unstructured data processing with best practices.^27^ Finally, independent agencies or governing bodies were proposed to oversee and ensure safe data sharing, preservation of intellectual property and valid applications.^18,19^

## Additional Recommendations

During the literature review, we identified two additional, overarching recommendations for unstructured data use, which are described subsequently.

### Collaborations with all stakeholders

Several sources stressed the importance of stakeholder collaboration in health research when combining different data sources for knowledge enrichment.^20,27,28^ The inclusion of with public and patient advocacy groups and other relevant stakeholders was highly recommended^20^ to ensure wide public acceptance and patient trust.^21,35^ Broad stakeholder involvement was also seen as crucial to increase data sharing and to minimize wasted efforts from research study duplication.^27^ Collaborative efforts among academic and commercial organizations (e.g., digital device or sensor manufacturers) can facilitate large-scale data integration and create synergies.^20,28^ Stakeholder and patient engagement during in the unstructured data integration, analysis, and interpretation provides relevant context and feedback on the meaningfulness of results.^20,27^

### Documentation and transparency

Proper documentation and transparency during the entire data flow were repeatedly mentioned as essential steps to achieve reliability, replicability, reproducibility and validity of studies, as well as facilitating the standardization efforts.^17,24,27,28^ The EMA framework emphasizes documentation as an important means to achieve reliability, repeatability, accuracy, clinical validity, generalizability, and clinical applicability of the novel methodologies.^27^ In the context of digital health technologies, United States Food and Drug Administration (FDA) recommendations suggest documenting the device and algorithm input and output, and to provide plans for data loss minimization, missing data handling, or patient inclusion for results. Furthermore, the FDA recommendations call for transparency of all processing steps from raw data to algorithm and at all data workflow stages.^27^ Transparency regarding the analysis process can also assist with the assessment of whether study findings were clinically significant.^24^ Specifically, studies relying on large databases will produce many statistically significant, but clinically meaningless results. This “overpowering” of statistical tests by large sample sizes should be made transparent through reporting of effect size determinants and complementation by clinical interpretation.^17^

#### Proposal for a feasibility and planning checklist for unstructured data enrichment

Many studies highlighted the need for further research and guideline development on best practices to use and integrate unstructured data in health research.^18,20,21,22,23,28,29^ In **Table 1**, we provide a set of guiding questions to inform early study planning and the assessment of the feasibility of studies. These questions are based on the described challenge areas, which have been expanded to align with the breadth of proposed solutions from our review.

**Table 1.**
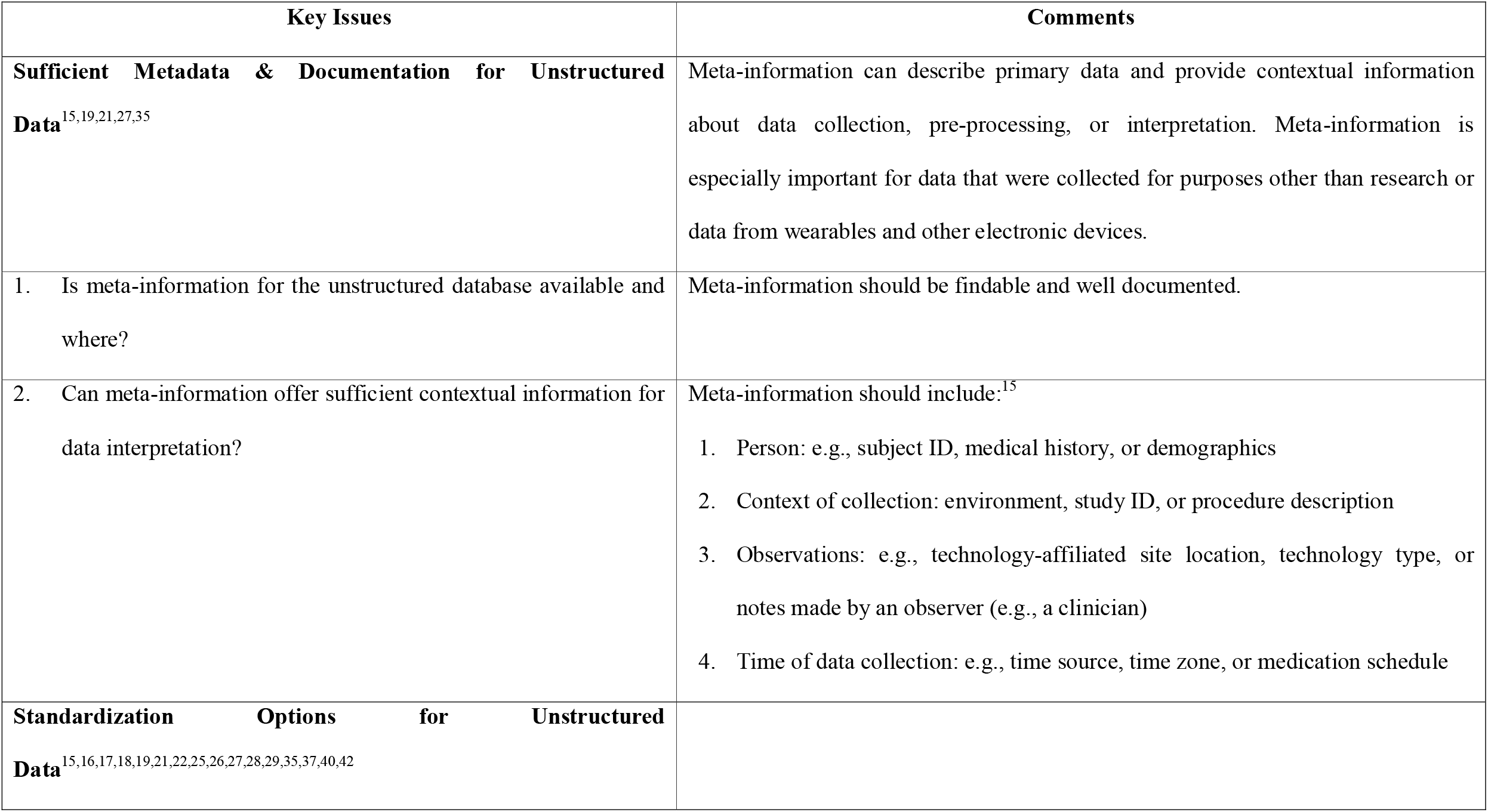
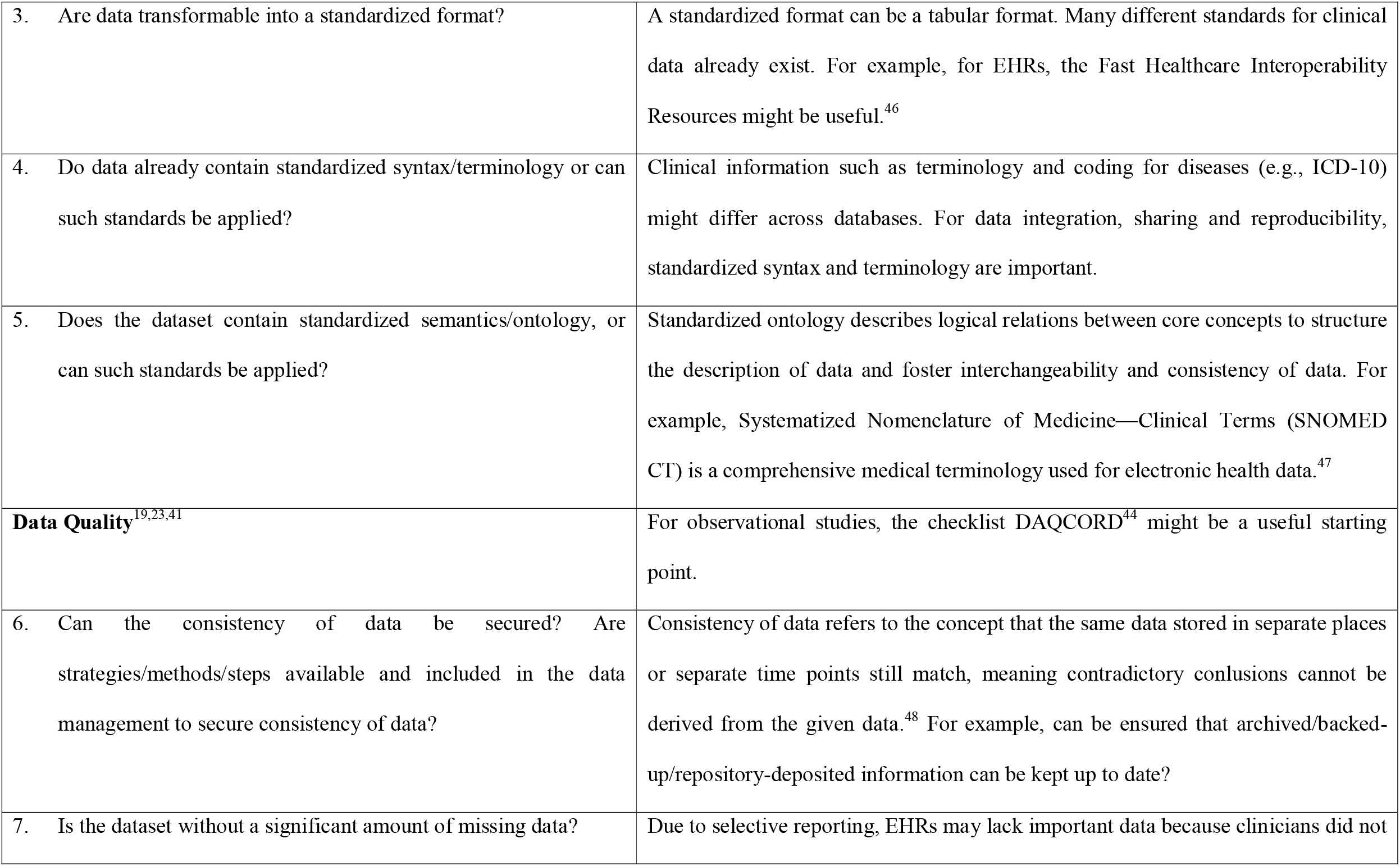
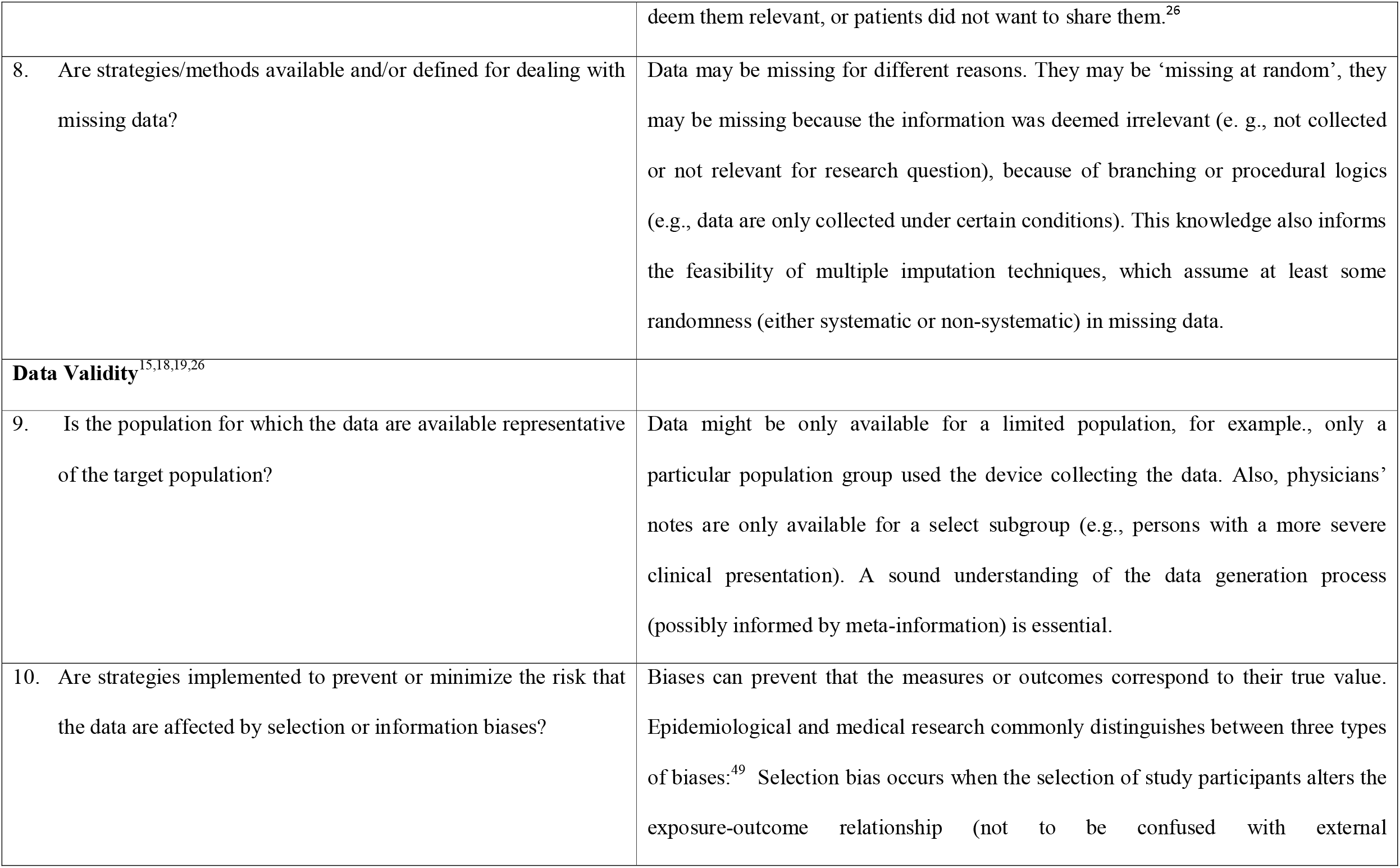
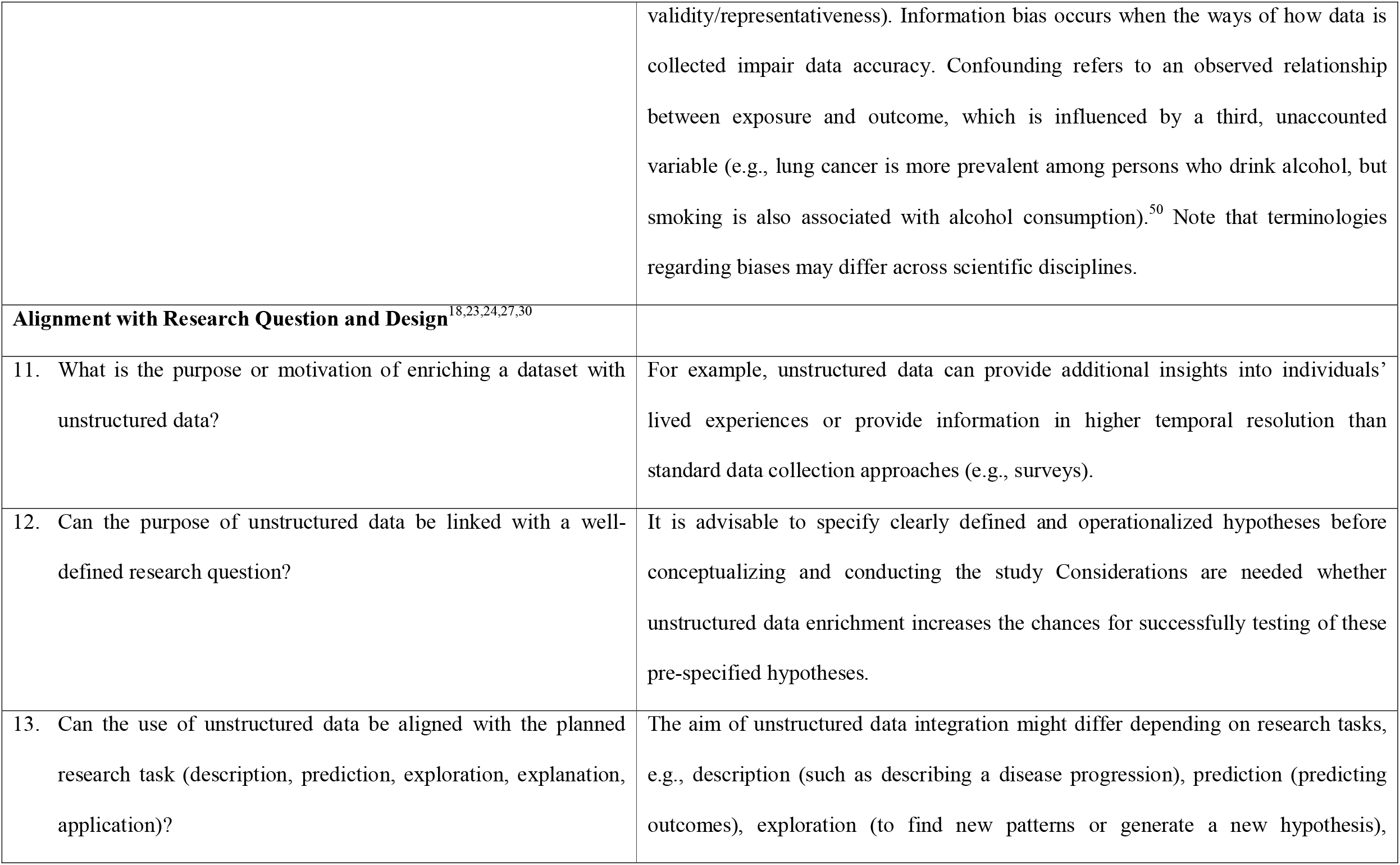
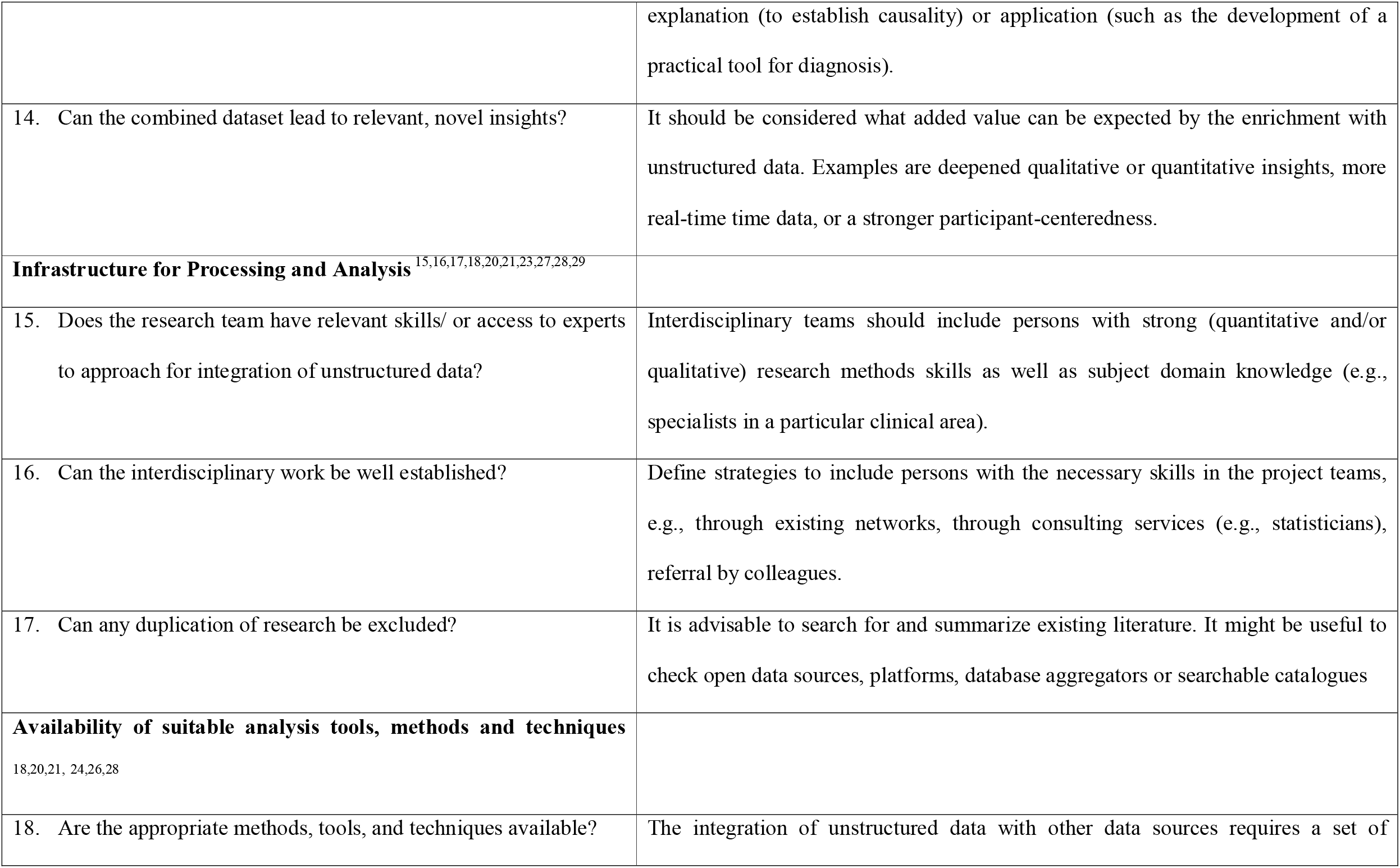
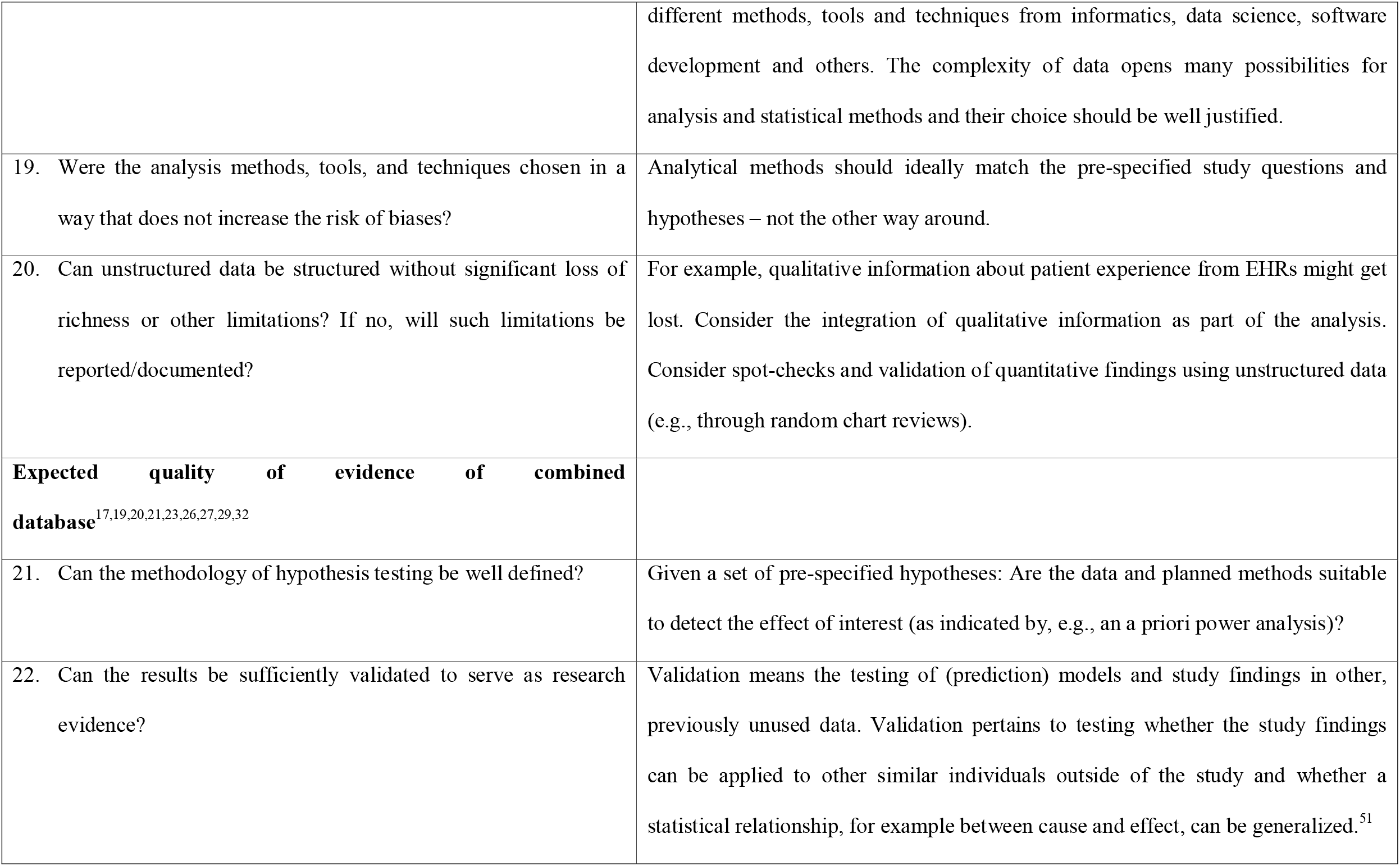
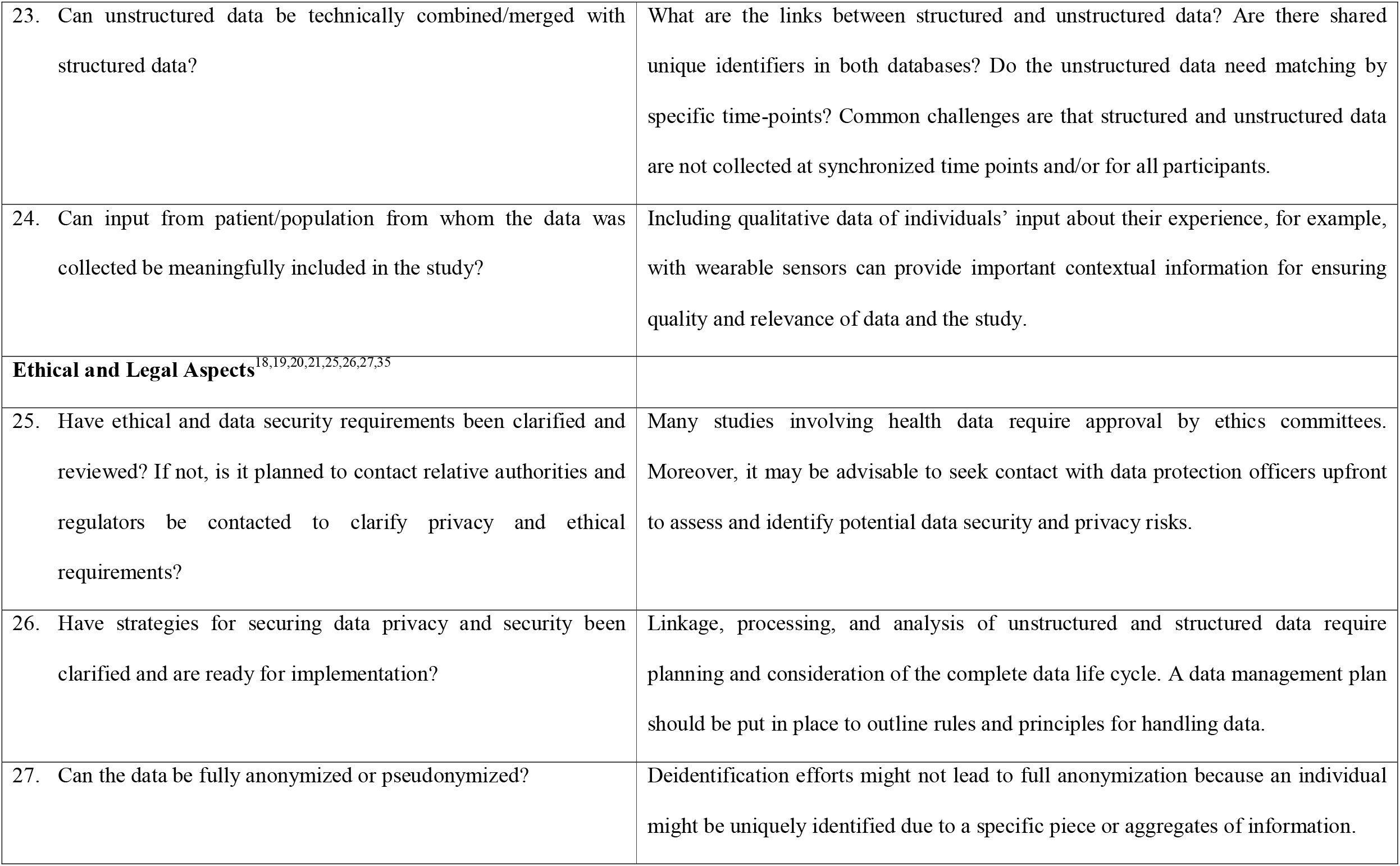
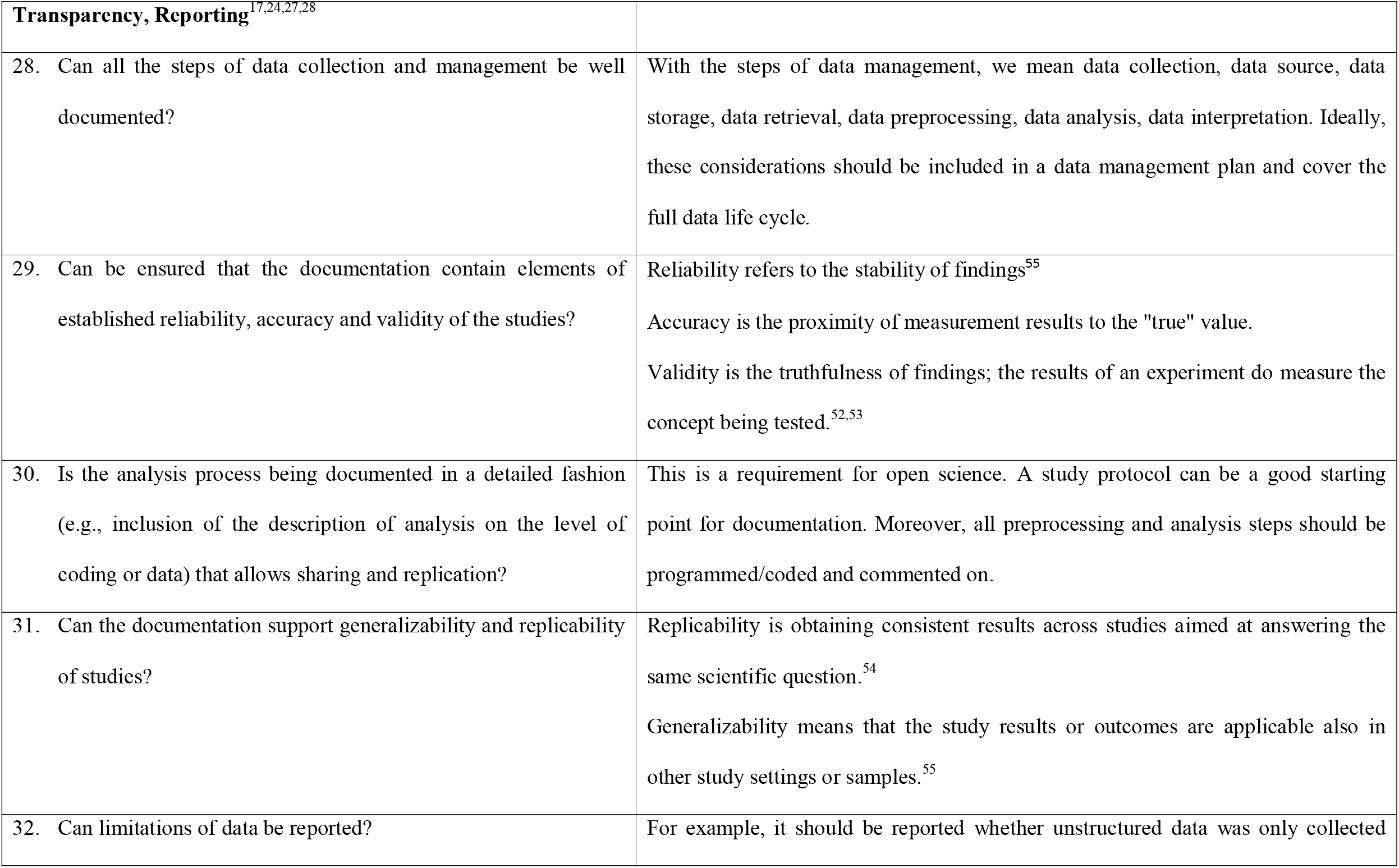
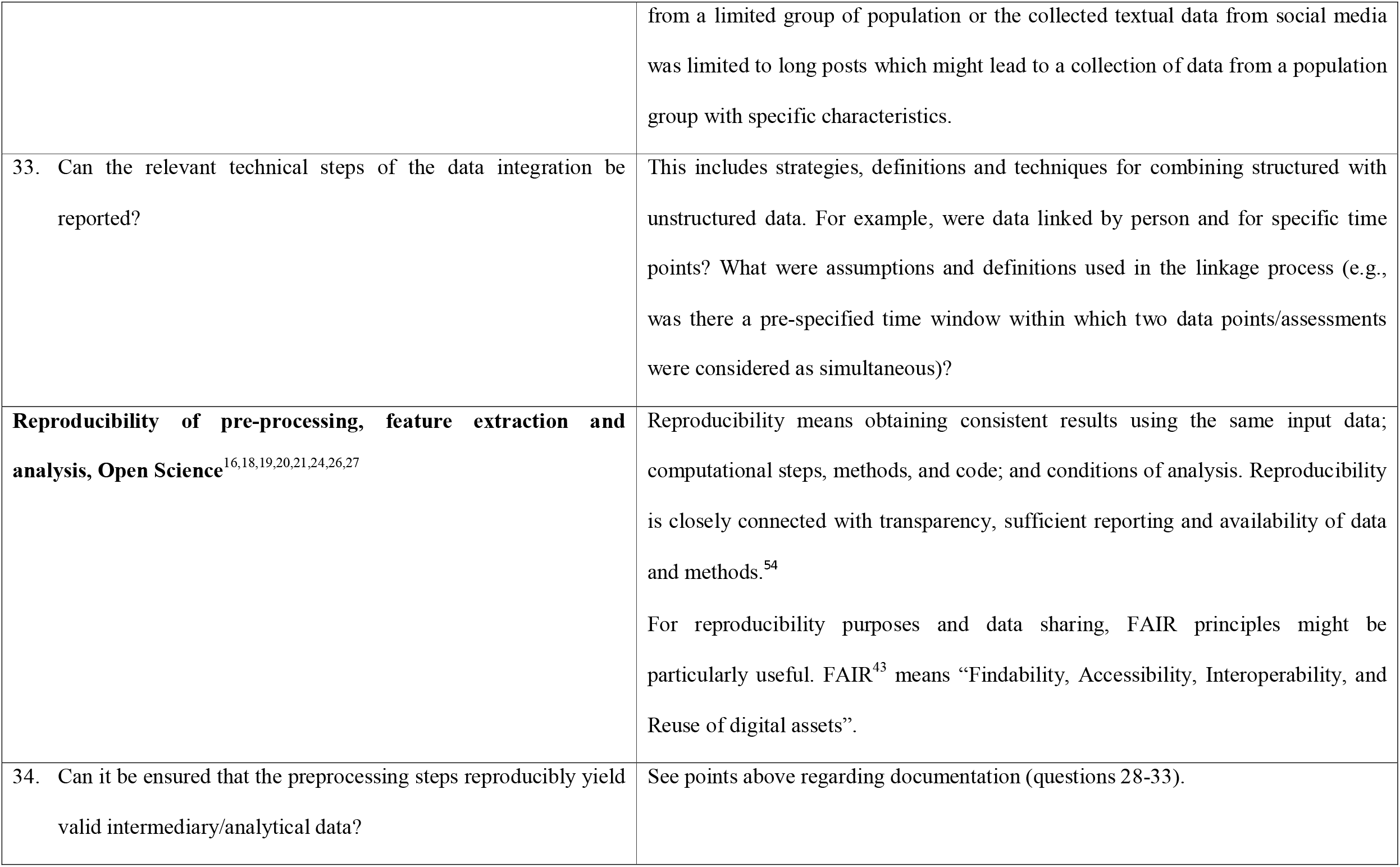
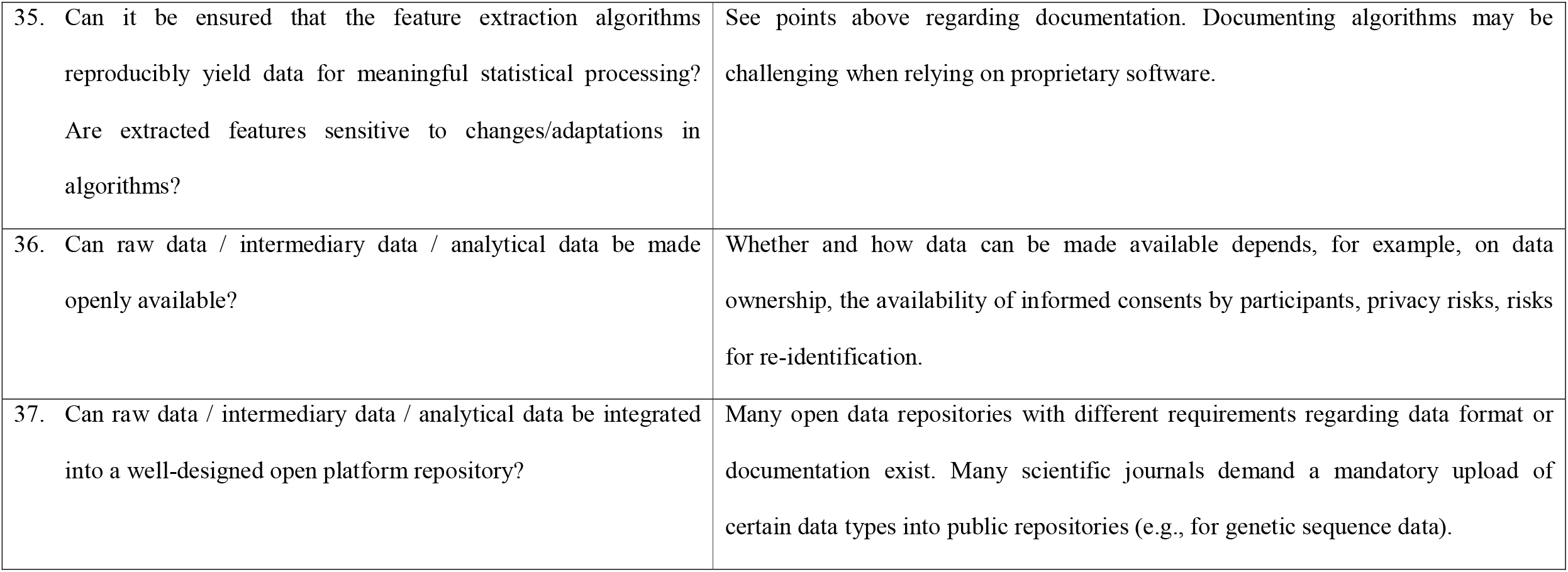
The checklist for early study planning and the assessment of the feasibility of studies using digital unstructured data.

## Discussion

### Summary of findings

Our systematic narrative review provides an overview of challenges and best practices associated with the combination of unstructured data with other data sources in health research, which we refer to as digital unstructured data enrichment. In our review, we identified seven prevalent challenge areas in enabling digital unstructured data enrichment: 1) the lack of meta-information for unstructured data, 2) standardization issues, 3) data quality and bias in unstructured data, 4) infrastructure and human resources, 5) finding suitable analysis tools, methods and techniques, 6) alignment of unstructured data with a research question and design, as well as 7) legal and ethical issues. For each challenge area, we summarized proposed possible solutions together with two additional recommendations that span across all challenge areas. We also summarized literature and experience-based checklist questions to inform initial study planning about the feasibility of research studies aiming to complement existing health data with digital unstructured data.

### Description of main requirements and solutions to enable unstructured digital data enrichment

All our studies revealed challenges associated with the digital unstructured data enrichment in health research, many of which might endanger scientific rigor and quality of health studies. For example, the frequently unclear suitability of digital unstructured data to address concrete research questions or allow for proper research study design^18,21,26^ may lead to possible biases, threatening the external and internal validity of studies. The validity of studies might also be endangered by applying not suitable analytical tools and methods. Furthermore, the findings of the study may lack generalizability limiting its use to specific research tasks and questions (e.g., hypothesis-generation).^17,18,21,26^ The lack of meta-information might hinder a proper interpretation of the data and consequently limit their use for enrichment purposes. Further problems are that the data can be placed so centrally that any bias will be strongly reflected in the results, also in the formation of data-driven categories. The most discussed challenge of standardization issues might hinder replicability and generalizability of research studies. Finally, ethical and legal issues, such as the risk of patient re-identification when disparate data sources are combined, pose additional challenges to digital unstructured data enrichment.

While many of the challenges to enable digital unstructured data enrichment are not specific to the use of unstructured data and are well known (e.g., data quality or standardization issues), other challenges, such as difficulties to align data with research questions or challenges pertaining to special skills or infrastructure needs, may be aggravated with the use of unstructured data due to their complexity. One of the key challenges might be the lack of open and collaborative platforms that can foster not only joint standardization but also validation efforts.^15,21,28^ Oftentimes, the attractive characteristics of unstructured data that might add value to research are the ones that pose the most challenges. The data granularity and large, often international, population-based sample can enhance disease understanding or monitoring but also lead to methodological challenges, for example, regarding validity and choice of tools for analyses.^24,26^

The possible solutions and additional recommendations are important to sustain interchangeability, validity, reliability, generalizability, and reproducibility of studies. The review revealed that the possible solutions are less frequently and systematically discussed than the challenges. Several sources discussed challenges without referring to the existing solutions or offering new proposals for possible solutions. The complexity of the digital unstructured data enrichment is also reflected in the possible solutions that can address several challenge areas. The interdisciplinary collaboration, open science and transparency were one of the most requested possible solutions.

### Requirement for guidance on digital unstructured data enrichment

Our review also revealed that despite the wide usage of unstructured data in health research and discussed challenges, there is a lack of a systematic approach and guidelines for researchers to address the observed challenges. Several of the selected articles acknowledged the need for more guidance,^18,20,21,22,23,28,29^ oversight or monitoring from agencies^18,27,42^ and interdisciplinary teamwork and exchange to establish methodological approaches in the context of utilizing unstructured data in health research.^21,26^ Only a few studies directly mentioned existing frameworks and standards such as EMA recommendations,^27^ FAIR principles,^28^ openHR^21^ or DAQORD framework^19^ in this context. Recent efforts to provide guidelines are mainly focused either on a specific type of unstructured data or specific challenges, for example, guidelines and standards for the use of social media data,^56,57^ guidelines regarding the use of EHRs,^58,59^ checklists and frameworks for evaluating the measurements made by digital technologies^60^ or algorithms used for data analysis.^61,62^ However, it is up for discussion whether these particular frameworks and guidelines are suitable to provide general guidance on challenges connected with digital unstructured data enrichment.

Our findings also reveal an underreporting of information relevant to digital unstructured data enrichment in health research. For example, current reporting guidelines such as STROBE^63^ do not cover unstructured data-enriched analyses. In the assessed studies, challenges relevant to integrating unstructured data with other health data sources were rarely mentioned. Our studies usually provided description of data collection and preprocessing and addressed issues of noisy and missing data. However, they often lacked a description of data limitations or strategies how to ensure data quality. In light of the growing volume and importance of unstructured data in health research, experience sharing should be increasingly encouraged – either in published literature (e.g., also in appendices) or in other outlets. The lack of reporting and unavailability of guidelines not only hampers study reproducibility but also presents missed opportunities for learning and capacity building.

All this points to a growing need to define systematic ways of how to approach digital unstructured data enrichment in health research. The numerous challenges directly linked with unstructured data use or digital unstructured data enrichment should be reflected in a systemic guidance on how to properly integrate digital unstructured data in health research. Our review identified a special need for guidance to establish common standards to enable digital unstructured data enrichment to help researchers in the first stages of study planning and to assess the feasibility of studies integrating unstructured data.^16,18,27^ The checklist derived from our review provides a first, pragmatic step towards classifying challenges and developing methodologies in health research involving digital unstructured data enrichment. In next steps, we hope to encourage specific research fields to dive deeper into our proposed checklist and adapt it to terminologies and issues that might be of a greater relevance in their respective research fields.

### Limitations

Although based on a systematic search and extraction process, we restricted our search to a few specific research fields due to the prevalent and growing use of unstructured data in these fields. Furthermore, we did not include books and book chapters. Therefore, our overview is likely not comprehensive. Furthermore, we a priori excluded imaging data and bioinformatics data from our literature search, which are an important source of unstructured data, but are often analyzed with highly specialized tools. In the systematic narrative review, we did not specifically discuss challenges and obstacles that are linked with learning algorithms used for unstructured data integration or data analysis/interpretation. However, there is also precaution and guidance needed for choices about learning algorithms. Machine learning and deep learning algorithms are not immune to errors, biases and other limitations that can negatively impact validity, objectivity, and reproducibility of studies.

## Conclusion

The integration of unstructured data into structured databases opens new avenues for more person-centered, contextualized, or more real-time analyses. However, multiple methodological and conceptual challenges demand attention, ideally even before an analysis is undertaken. A clear definition and focus on suitable study questions, interdisciplinary team-work, or transparent documentation and open science are key ingredients towards a more robust unstructured data enrichment methodology. Overall, our review also points to a need of more guidance – and possibly also standards for reporting results of digital unstructured data studies. Awareness should be raised among researchers to openly document encountered challenges and possible solutions in unstructured data enrichment projects to enable experience exchanges and learning. Moreover, existing reporting guidelines such as STROBE should consider adding specific instructions on the documentation of unstructured data enrichment processes.

## Data Availability

All data produced in the present study are available upon reasonable request to the authors

## Data Availability

The data from the papers that support the findings of this study are publicly available. All data used in this review can be provided upon request.

## Author contributions

The review was conducted as a collaborative effort in the Health Community of Digital Society Initiative at the University Zurich (the members’ list is to be found in the **Appendix 3**). The core research team (consisting of JS, PD, VvW) defined the initial plan for the review together with the research question and search syntax as well as conducted the literature search, screening steps, data extraction, data synthesis and wrote and revised the manuscript. MS, CH, CS assisted with the initial plan for the review, conducted the literature search, helped interpret the findings and revised the final manuscript. The entire process was discussed in the Health Community meetings. In addition, every step was consolidated, evaluated and approved by the Executive Committee (consisting of AHW, MW, GS, OG, DAE, KS) that provided further insights, comments and contribution to the manuscript.

## Ethics Declaration

The authors declare no competing interests.

**Correspondence** should be addressed to Viktor von Wyl.

