## Appendix 3 for "Challenges and best practices for digital unstructured data enrichment in health research: a systematic narrative review"

**Appendix 3. The members' list of Health Community of Digital Society Initiative, University of Zurich, Zurich, Switzerland**

Mathias Allemand, Fabio Balli, Michael Baudis, Patrich Beeler, Jürgen Bernard, Walter Bierbauer, Nikola Biller-Andorno, Mathias Bonmarin, Markus Christen, Paola Daniore, Noemi Dannecker, Burcu Demiray, Mateusz Dolata, Tilia Ellendorff, Dominik Ettlin, Andri Färber, Joël Floris, Thomas Friemel, Philipp Fürnstahl, Sarah Geber, Federico Germani, Daniel Gero, Arko Ghosh, Marcin Gil, Felix Gille, Andrea Glaessel, Oliver Grübner, Christina Haag, Eva Maria Hakanson, Andrea Horn, Gioacomo Invideri, Jeffrey Iqbal, Philipp Kerksieck, Viktor Kölzer, Michael Krauthammer, Tobias Kowatsch, Onicio Leal-Neto, Soeren Lienkamp, Minxia Luo, Anke Maatz, Ivor Mardesic, Mike Martin, Julian Mausbach, Gianluca Miscione, Corine Mouton Dorey, Margot Muetsch, Pashalis Naoumis, Vasileios Nittas, Farhad Nooralahzadeh, Kimon Papadopoulos, Milo Puhan, Fabio Rinaldi, Sonja Schläpfer, Gerold Schneider, Urte Scholz, Gerhard Schwabe, Bettina Friederike Schwind, Jana Sedlakova, Chloé Sieber, Giovanni Spitale, Mina Stanikic, Kaspar Staub, Nina Steinemann, Jürg Streuli, Rasita Vinay, Viktor von Wyl, Ning Wang, Fabian Winiger, Claudia Witt, Markus Wolf, Aleksandra Zumbrunn
