## Appendix 1 for "Challenges and best practices for digital unstructured data enrichment in health research: a systematic narrative review"

**Appendix 1. Search Syntax**

In Title/Abstract:

(“unstructured data” OR “big data” OR “multimodal data” OR “text data” OR “textual data” OR “voice data” OR “sensor data” OR “video data” OR “social media data” OR “speech data” OR “wearable data” OR “sensing data”) AND (“health*” OR “medical” OR “clinical” OR “patient” OR “mHealth” OR “digital health” OR “patient-generated” OR “PGHD” OR “patient-reported”)

All fields:

(“cardiovascular” OR “atrial fibr*” OR “myocardial infarction” OR “cardiolog*” OR “depression” OR “anxiety” OR “mental health” OR “mental disorder*” OR “neurolog*” OR “multiple sclerosis” OR “MS”) AND (“integrat*” OR “appl*” OR “enrich*” OR “combin*” OR “harmoniz*” OR “synth*” OR “adopt*” OR “link*” OR “fusion”) AND (“problem” OR “challeng*” OR “difficult*” OR “obstacle” OR “gap” OR “need” OR “issue*” OR “barrier*” OR “requirement*” OR “limitation*” OR “incompatib*”)
