## Appendix 2 for "Challenges and best practices for digital unstructured data enrichment in health research: a systematic narrative review"

**Appendix 2. Supplementary Table 1. Description of the included studies**

| First author, year of publication | Type of paper | Field (subfield) | Motivation for using unstructured data | Type of unstructured and structured data being integrated |
|---|---|---|---|---|
| Badawy R., 2019**Error! Bookmark not defined.** | General (Review) | Neurology (Parkinson's disease) | • Information enrichment: insights into patients' symptoms, disease progression, and treatment efficacy<br>• Cost effective and efficient<br>• Remote, long-term monitoring in naturalistic settings<br>• Increased statistical power of clinical trials<br>• Reduction random sampling issues | Data from digital health technologies; focus on metadata |
| Baldassano S. N., 2019**Error! Bookmark not defined.** | General (Review) | Neurology (Status Epilepticus) | • Personalized medicine<br>• Patients' stratification for prognostication<br>• Patients' identification for interventions | Big data, more specifically: EHRs, administrative databases (e.g., data collected for billing purposes), iEEG, multimodal ICU data, physiological data (e.g., heart rate, respiratory rate) |
| Blair L. M., 2016**Error! Bookmark not defined.** | General (Overview) | Mental Health (Pediatrics) | • Ease of access<br>• Nationally representative sampling<br>• Efficiency and lower cost | Publicly available big data |
| Espay A. J., 2016**Error! Bookmark not defined.** | General (Review) | Neurology (Parkinson's Disease) | • Information enrichment<br>• Treatment optimization<br>• Maximization of "ecological" validity<br>• Monitoring<br>• High resolution descriptions<br>• Quality of care<br>• Personalized medicine | Data from health technology (e.g., wearable devices) |

| | | | | |
|---|---|---|---|---|
| Foreman B., 2020**Error! Bookmark not defined.** | General (Review) | Neurology (Neurocriticial Care) | • Patient cohorts' enrichment<br>• Population-level insights<br>• Precision medicine<br>• Evaluation of interventions<br>• Cost efficient<br>• Precision medicine | ICU data, EEG, unstructured free-text clinician notes, Digital Imaging and Communications in Medicine standard imaging, Logical Observation Identifiers Names and Codes (LOINC), standard laboratory values |
| Hafferty J. D., 2017**Error! Bookmark not defined.** | General (Commentary) | Mental Health | • Naturalistic clinical settings<br>• Analysis of rarer clinical conditions or subject areas<br>• Prediction<br>• Personalised medicine research<br>• Better modelling of the 'bio-psycho-social' outcomes of psychiatric illness | Big Data in general |
| Hemingway H., 2017**Error! Bookmark not defined.** | General (Overview) | Cardiology | • High resolution results and large-scale studies<br>• Real time analytics<br>• Treatment improvement<br>• More efficient and cost-effective methods | EHRs (structured and unstructured data); biobanks, genomic consortia; any researcher-generated data (e.g., omics data) |
| Rodriguez A., 2018**Error! Bookmark not defined.** | General (Overview) | Neurology (Neurocritical care) | • Better patients' stratification and management<br>• High resolution descriptions | Genomic, imaging, physiological, and phenotypic<br><br>information (e.g., EEG), pre- and post-psychometric testing, EHR |
| Rumsfeld J. S., 2016**Error! Bookmark not defined.** | General (Review) | Cardiology | • Prediction<br>• Disease detection<br>• Monitoring<br>• Providing high resolution descriptions and large-scale studies<br>• Prescriptive analytics<br>• Quality of care and performance measures<br>• Public health improvement | Big Data in general |

| | | | | |
|---|---|---|---|---|
| Schofield P., 2017**Error! Bookmark not defined.** | General (Editorial) | Mental Health | • Information enrichment: data about hard-to-reach groups, new relations between clinical outcomes and other factors | EHRs, data from other databases (e.g., hospital episode statistics, educational data) |
| Shen B., 2018**Error! Bookmark not defined.** | General (Review) | Neurology (Parkinson's disease) | • Screening<br>• Holistic descriptions<br>• Information enrichment | EHR, physiological data (EEG, electrocardiogram), omics data, neuroimaging data, epidemiological data |
| Silverio A. 2019**Error! Bookmark not defined.** | General (Overview) | Cardiology | • Insights into patients' life, behavior, and symptoms<br>• High resolution descriptions and large-scale studies<br>• Monitoring<br>• Public health improvement<br>• Prediction<br>• Personalized medicine<br>• Quality of care and performance measures<br>• Genetic insights | 'Big Data' in general |
| Stephenson D., 2020**Error! Bookmark not defined.** | General (Review) | Neurology (Parkinson disease) | • Evaluation and subsequent approval of novel treatments<br>• Assessment of aspects of the disease<br>• Improvement of outcome measures<br>• Better patient enrolment and stratification<br>• Monitoring<br>• Real-world data<br>• Implementation of remote trials | Data from broad range of digital health technologies (e.g., smartphone applications, wearable sensors, GPS, EEG, digital diaries) |
| Termine A., 2021**Error! Bookmark not defined.** | General (Review) | Neurology (neurodegenerative disorders such as Alzheimer's and | • High resolution descriptions<br>• Accuracy of analytical results<br>• Quality of care<br>• Prediction<br>• Personalized and precision medicine<br>• More efficient and cost-effective | EHRs, multi-omics, neuroimaging and wearable sensors data |

| | | | | |
|---|---|---|---|---|
| | | Parkinson's diseases) | | |
| van den Heuvel L., 2020**Error! Bookmark not defined.** | General | Neurology (Parkinson's disease) | • Information enrichment<br>• Real-time data<br>• Personalized precision medicine and decision-making process | Big data in general |
| Andy A. U., 2021**Error! Bookmark not defined.** | Research article | Cardiology (atherosclerotic cardiovascular disease) | • Prediction of individual risks<br>• Better understanding of individual risks<br>• Precision medicine | Facebook status updates, medical record data |
| Clark R. A., 2019**Error! Bookmark not defined.** | Research article | Cardiology (Acute coronary syndrome) | • Real world assessment | Quantitative and digital data<br><br>collection (GPS and GIS), qualitative patient-reported experiences (satisfaction survey), a<br><br>discharge education recall questionnaire |
| Haines-Delmont A., 2020**Error! Bookmark not defined.** | Research article | Mental Health (Suicide Prevention) | • Exploration<br>• Prediction<br>• Real-time, context-related monitoring<br>• Ecologically momentary assessment in naturalistic settings<br>• Real-time classification<br>• Quantification of human behavior | Facebook, user data, clinician ratings, passive sensor data |
| Jacobson N.C., 2020**Error! Bookmark not defined.** | Research article | Mental Health (Social Anxiety) | • Prediction<br>• Monitoring | Social Interaction Anxiety<br><br>Scale; Depression, Anxiety, and Stress Scale; Positive Affect Negative Affect Schedule; passive sensor data |

| | | | | |
|---|---|---|---|---|
| | | | | (accelerometer data), incoming and outgoing calls and text timestamps |
| Li B., 2019**Error! Bookmark not defined.** | Research article | Cardiology (cardiovascular disease) | • Disease diagnostic<br>• Early-risk prediction | Medical big data, medical diagnostic records |
| Papadopoulos A., 2020**Error! Bookmark not defined.** | Research article | Neurology (Parkinson's disease) | • Unobtrusive, remote detection of early symptoms<br>• Real-time data | Passively captured data from smartphones: the tri-axial<br><br>acceleration values obtained from the Inertial Measurement Unit sensor of the smartphone and the keystroke timing data (press and release timestamps of each keystroke captured during typing with the smartphone's virtual keyboard) |
| Payrovnaziri S. N., 2019**Error! Bookmark not defined.** | Research article | Cardiology (Acute myocardial infarction, post myocardial infarction syndrome) | • Prediction<br>• Effective and efficient<br>• Precision medicine<br>• Quality of care<br>• High resolution descriptions | Demographic and admission data (from EHR), free text from discharge summary |
| Ross E. G., 2019**Error! Bookmark not defined.** | Research article | Cardiology (periphery artery disease) | • Risk prediction of local populations or more specific disease states<br>• Automatization of risk stratification | International Classification<br><br>of Diseases, Ninth Revision (ICD-9) codes, Current Procedural<br><br>Terminology codes, lab test values, prescription medications,<br><br>vital signs, unstructured clinical notes |

| | | | | |
|---|---|---|---|---|
| Sajal S. R., 2020**Error! Bookmark not defined.** | Research article | Neurology (Parkinson's diseases) | • Increased accuracy<br>• Understanding of disease history, progression, and risk factors | Rest tremor and vowel phonation data acquired by smartphones |
| Sükei E., 2021**Error! Bookmark not defined.** | Research article | Mental health | • Prediction of mood states<br>• Assessment of behavioral features | Passively sensed behavioral data from 6 sources, users' record information |
| Ahn I., 2021**Error! Bookmark not defined.** | Paper on Databases | Cardiology | • Early-risk prediction<br>• Discovery of risk factors and detection of their interactions<br>• Prevention<br>• Improvement of treatment planning | Structured data from electronic health records: demographics, vital signs, medication, laboratory test results, patient history questionnaire. Unstructured data from text readings of electrocardiogram, coronary artery computed tomography, single-photon emission computed tomography |
| Matoba T., 2018**Error! Bookmark not defined.** | Paper on Databases | Cardiology | • Real-time data<br>• Precision medicine<br>• Complementation to randomized clinical trials | Medical records, electronic<br><br>data regarding medical activity and procedures; key data regarding coronary angiography and percutaneous coronary intervention |
| Perera G., 2016**Error! Bookmark not defined.** | Paper on Databases | Mental health | • Information enrichment<br>• Novel investigations<br>• Detection of patterns in patient care and treatment habits<br>• Enhancement of research question | EHRs, diverse databases |

**Appendix 2. Supplementary Table 2. Description of Challenge Areas.**

| Challenge area | Definition/Description | Relevance |
|---|---|---|
| **1. Lack of meta-information for unstructured data** | All topics related to documentation of size, content, context, and format of unstructured data are included in this challenge area. | - To *scrutinize* and possibly *avoid biased* assumptions about the unstructured data.**Error! Bookmark not defined.**<br>- To provide *contextual information* for data interpretation**Error! Bookmark not defined.**,**Error! Bookmark not defined.** (e.g., when and where data from wearable sensors were generated). The contextual information can enable a more *robust analysis* and *meaningful data interpretation**Error! Bookmark not defined.*** which is important to *evaluate* the *accuracy* of the unstructured data *and* its *interpretability*.**Error! Bookmark not defined.**<br>- To facilitate the evaluation of the *quality and reliability* of the unstructured data.**Error! Bookmark not defined.**<br>- To facilitate replicability of studies.<br>- To facilitate the *interchangeability and reuse* of unstructured data.**Error! Bookmark not defined.**,**Error! Bookmark not defined.**<br>- To facilitate the process of *validation for regulatory acceptance* (e.g., in the context of digital health technologies).**Error! Bookmark not defined.** |
| 2. **Standardization Issues** | All challenges related to the conversion of unstructured data into a common standardized format that enables it to get shared, linked and used across different settings are included in this challenge area. | - To facilitate/foster effective data access, reuse of data for research projects and ultimately interoperability, interchangeability, and linkage of data.**Error! Bookmark not defined.**,**Error! Bookmark not defined.**,**Error! Bookmark not defined.**,**Error! Bookmark not defined.**<br>- To avoid duplication of research.**Error! Bookmark not defined.**<br>- To facilitate data interpretation and extraction of correct information.**Error! Bookmark not defined.**,**Error! Bookmark not defined.**,**Error! Bookmark not defined.**,**Error! Bookmark not defined.**,**Error! Bookmark not defined.**<br>- To enable data consistency.**Error! Bookmark not defined.** |

| | | |
|---|---|---|
| **3. Data Quality and Bias in Data** | All topics concerning data accuracy, reliability, validity, and consistency are included in this challenge area. | - To increase/secure data accuracy, reliability, validity, reproducibility, replicability and consistency.<br>- To avoid a loss of efficiency in achieving study goals, errors in analysis.<br>- To facilitate interpretation of findings. |
| **4. Infrastructure** | All topics related to IT infrastructure that enable or facilitate data management, access, sharing, and processing are included in this challenge area. | - To facilitate access to data that researchers need for their research.**Error! Bookmark not defined.**,**Error! Bookmark not defined.**<br>- To avoid missed opportunities by lack of accessibility to relevant data sources.<br>- To reduce research costs.**Error! Bookmark not defined.**,**Error! Bookmark not defined.,Error! Bookmark not defined.** |
| **5. Finding suitable analysis tools, methods, and techniques** | All topics related to the methodological choices of how unstructured data are processed and analyzed are included in this challenge area. | - To facilitate the complex process of cleaning and analyses of large and complex datasets.**Error! Bookmark not defined.**,**Error! Bookmark not defined.,Error! Bookmark not defined.,Error! Bookmark not defined.,Error! Bookmark not defined.,Error! Bookmark not defined.,Error! Bookmark not defined.**<br>- To decrease risk for bias in research.**Error! Bookmark not defined.**,**Error! Bookmark not defined.**<br>- To increase/secure data accuracy, reliability, validity, reproducibility, replicability and consistency. |
| **6. Alignment with a research design and/or research question** | All topics related to the broader theoretical issues of how the use and/or integration of unstructured data is linked with an appropriate research design and question is included in this challenge area. | - To ensure scientific rigor and validity.**Error! Bookmark not defined.**<br>- To determine the most suitable data analysis approach.**Error! Bookmark not defined.** |
| **7. Ethics & Legal Issues** | All topics emerging from ethical and legal concerns or risks either on the societal or individual level – such as privacy, confidentiality, safety, and discrimination – are included in this challenge area. | - Adherence to ethical and legal frameworks is a condition sine que non for research and requires no further justification.<br>- To facilitate a successful integration of unstructured data in health research as it can increase public trust and acceptance which might lead to increased availability of data sources. |
